# Urinary Metabolite Signatures Stratify Survival in Malignant Mesothelioma: A Noninvasive Prognostic Tool

**DOI:** 10.1101/2025.09.23.25336488

**Authors:** Leila Toulabi, Joshua K. Stone, Mohammed Khan, Raffit Hassan, Curtis C. Harris, Daxesh P. Patel

## Abstract

Malignant mesothelioma is a rare, aggressive malignancy with limited treatment options and few robust prognostic biomarkers. Current blood-based assays (MESOMARK, fibulin-3, osteopontin) are constrained by inconsistent sensitivity and preanalytical instability. Here, we evaluated four urinary metabolites—creatine riboside (CR), N-acetylneuraminic acid (NANA), cortisol sulfate (CS), and 27-nor-5β-cholestane-3α,7α,12α,24,25-pentol (cholestane pentol, CP)—previously linked to prognosis in lung cancer and intrahepatic cholangiocarcinoma, in a clinically annotated mesothelioma cohort (n=95; NCT01950572) with paired RNA-sequencing data. All four metabolites correlated positively with a validated 48-gene poor-prognostic gene-expression signature, with a standardized composite score showing the strongest concordance (Spearman R=0.34, p=0.001). Overall survival declined progressively with the number of elevated metabolites, and patients with all four elevated had the poorest prognosis (log-rank p<0.0001). Stratification of the composite score by quartile and by median reproduced this graded effect (p<0.0001 for both), and associations were independent of age, sex, and disease site. Urinary metabolite profiling thus offers a simple, repeatable, noninvasive prognostic tool that complements existing blood-based assays and may support risk stratification, longitudinal monitoring, and clinical-trial enrichment in mesothelioma.

## 1. Introduction

Malignant mesothelioma is an uncommon but aggressive cancer arising from the mesothelial lining of the pleura, peritoneum, or pericardium and is strongly associated with prior asbestos exposure. Despite multimodal therapy combining surgery, chemotherapy, and immune-checkpoint inhibition, median survival rarely exceeds 12–18 months, and there remains no broadly reliable, noninvasive prognostic biomarker [1].

Current biomarkers are predominantly blood-based. MESOMARK, the only FDA-cleared mesothelioma assay, quantifies soluble mesothelin-related peptides and offers high specificity but limited sensitivity for early-stage disease [2]. Fibulin-3 and osteopontin show greater sensitivity in selected contexts but vary markedly between plasma and effusion samples and are subject to thrombin cleavage in serum, complicating interpretation [3,4]. These constraints highlight an unmet need for biomarkers that are reproducible, less affected by preanalytical variability, and ideally accessible through noninvasive collection.

Metabolomics captures the integrated downstream output of genetic, transcriptomic, host, and environmental factors. Urine is particularly attractive because it can be collected noninvasively, repeatedly, and at low cost, with fewer clotting and processing artifacts than blood. Four urinary metabolites—creatine riboside (CR), N-acetylneuraminic acid (NANA), cortisol sulfate (CS), and 27-nor-5β-cholestane-3α,7α,12α,24,25-pentol (cholestane pentol, CP)—have been validated as prognostic in lung cancer and intrahepatic cholangiocarcinoma, implicating arginine/creatine metabolism, sialic-acid biology, glucocorticoid recycling, and cholesterol metabolism [5,6]. Their reproducible prognostic value across tumor types suggests they may reflect fundamental aspects of cancer biology that extend beyond organ-specific contexts.

We hypothesized that these four metabolites would also be prognostic in mesothelioma, given shared hallmarks of metabolic reprogramming, inflammation, and aggressive clinical behavior. Here we report that elevated urinary concentrations of CR, NANA, CS, and CP correlate with a validated poor-prognostic gene-expression signature and stratify overall survival in a dose-dependent manner, supporting their potential as practical, clinically deployable biomarkers in mesothelioma.

## 2. Materials and methods

### 2.1 Patients and biospecimens

Biospecimens were obtained from the Tissue Procurement and Natural History Study of Patients with Malignant Mesothelioma (ClinicalTrials.gov: NCT01950572) under an NCI Institutional Review Board–approved protocol, in accordance with the International Conference on Harmonisation–Good Clinical Practice (ICH-GCP) guidelines and the Declaration of Helsinki. All participants provided written informed consent. Between September 2013 and November 2019, 425 patients with histologically confirmed mesothelioma (Board-Certified Anatomic Pathologists, NCI Laboratory of Pathology) were enrolled regardless of asbestos exposure, sex, age at diagnosis, or personal or family history of cancer. RNA-sequencing data were available for 100 patients; the present analysis focused on 95 patients with matched urine metabolomic and transcriptomic data (Supplementary Table S1). Detailed protocols are provided in the Supplementary materials.

### 2.2 Metabolite quantification

Urine extracts were prepared and analyzed by ultraperformance liquid chromatography–tandem mass spectrometry (UPLC-MS/MS) as previously described [5,8]. Concentrations of CR, NANA, CS, and CP were measured using a Waters XEVO G2 ESI QTOF mass spectrometer with authentic analytical standards. CR (m/z 264.1196, retention time [RT] 0.4 min) and CP (m/z 561.3435, RT 6.3 min) were measured in electrospray ionization (ESI) positive mode; NANA (m/z 308.0982, RT 0.4 min) and CS (m/z 441.162, RT 5.5 min) in ESI negative mode. Concentrations were calculated against calibration curves (MassLynx, Waters) and normalized to urinary creatinine. Patients with metabolite concentrations below the cohort median were classified as “low” and those at or above the median as “high” (medians: CR 1.987 µM, NANA 9.234 µM, CS 21.011 nM, CP 5.877 nM).

### 2.3 Gene-expression signature scoring

A previously validated 48-gene poor-prognostic mesothelioma signature [7] was scored per patient from matched RNA-sequencing data using the published gene list.

### 2.4 Composite metabolite score

Individual metabolite concentrations were z-transformed within the cohort, summed across the four metabolites, and averaged per patient to yield a composite score. Patients were stratified into quartiles (Sum_quad I–IV) and into low/high groups by median split (Sum_med) for survival analyses, and the composite score was also evaluated as a continuous variable.

### 2.5 Statistical analysis

All statistical analyses were performed in R 4.1.1. Group comparisons used the Mann–Whitney U test; correlations used Spearman’s rank coefficient. Overall survival was analyzed by the Kaplan–Meier method (log-rank test) using the survival and survminer packages, with verification in Stata. Two-sided p<0.05 was considered statistically significant.

## 3. Results and discussion

### 3.1 Cohort characteristics

Ninety-five mesothelioma patients with paired urine metabolomic and RNA-seq data were analyzed. The cohort was balanced for sex (54% male, 46% female) and disease site (49% pleural, 46% peritoneal), with 62% under 60 years of age and 55% reporting asbestos exposure (Supplementary Table S1).

### 3.2 Urinary metabolites correlate with a prognostic gene signature

Urinary CR, NANA, CS, and CP were each evaluated against the 48-gene poor-prognostic expression signature [7]. Spearman correlations were positive across all four metabolites (Fig. 1A–D); NANA, CS, and CP reached statistical significance (NANA R=0.25, p=0.014; CS R=0.30, p=0.0034; CP R=0.25, p=0.0017), and CR showed a concordant trend (R=0.19, p=0.067). When dichotomized at the median, patients in the “high” category had significantly higher gene-signature scores than those in the “low” category (Mann–Whitney p<0.05 for each metabolite). The composite metabolite score showed the strongest correlation (R=0.34, p=0.001; Fig. 1E), indicating that integrating information across metabolites improves concordance with the underlying molecular phenotype.

**Fig. 1.**
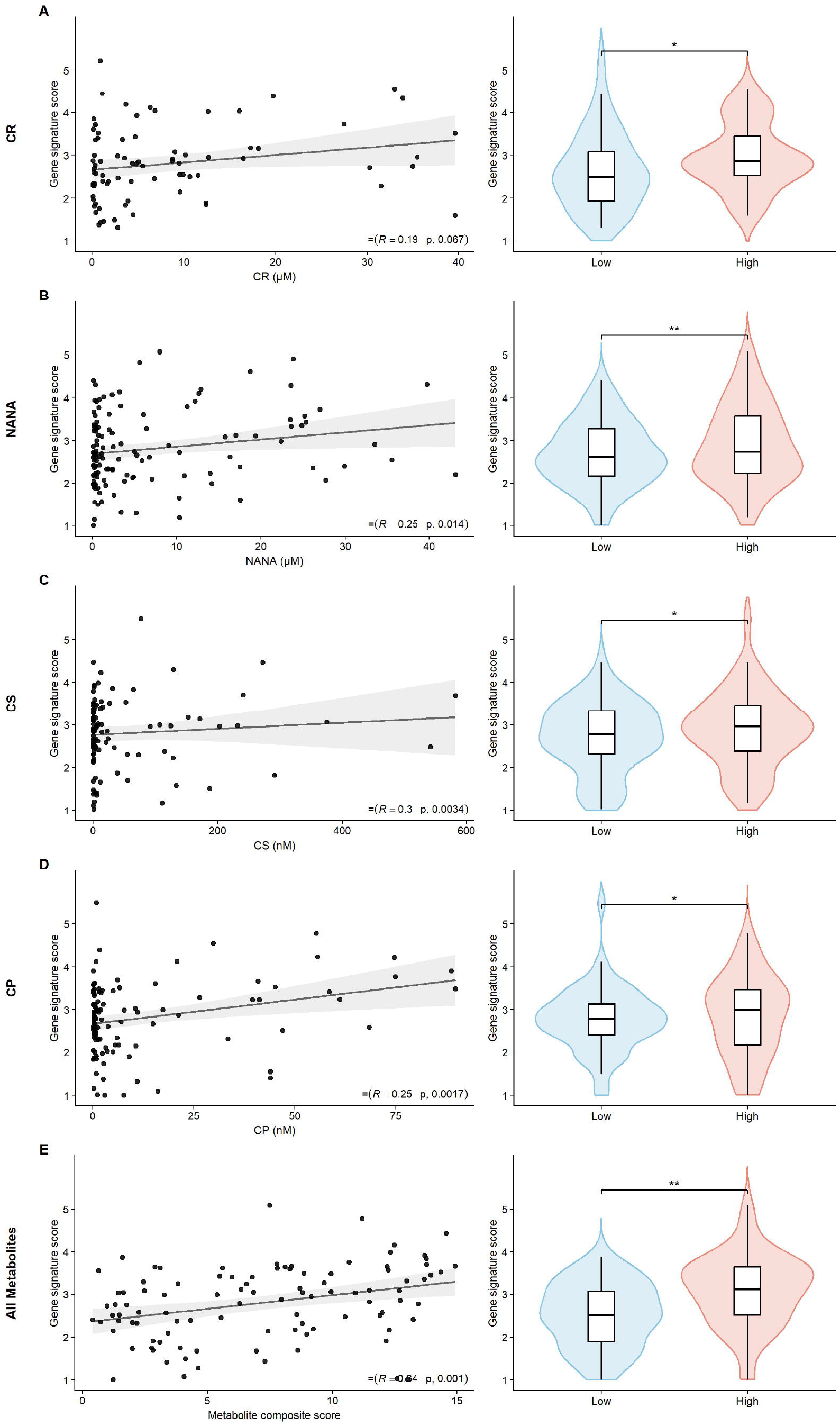
Urinary metabolite concentrations correlate with a validated 48-gene poor-prognostic expression signature in mesothelioma. (Left) Spearman correlation scatterplots between metabolite concentrations and 48-gene signature scores [7]. (Right) Patients were dichotomized at the median for each metabolite and compared by Mann–Whitney U test. (A) Creatine riboside, CR (low n=48, high n=47). (B) N-acetylneuraminic acid, NANA (low n=48, high n=47). (C) Cortisol sulfate, CS (low n=46, high n=47). (D) Cholestane pentol, CP (low n=46, high n=48). (E) Composite score across all four metabolites (low n=46, high n=47). ^*^p<0.05; ^**^p<0.01.

### 3.3 Metabolite levels are robust to clinical covariates

Metabolite concentrations did not differ significantly by age (<60 vs ≥60 years), pleural vs peritoneal site, or sex, with the exception of a modest CP elevation in female patients (Supplementary Figs. S1–S3). The independence of metabolite levels from these covariates supports their generalizability as prognostic biomarkers across patient subgroups and indicates that the prognostic signal reflects tumor biology rather than clinical heterogeneity.

### 3.4 Elevated urinary metabolites stratify overall survival

Kaplan–Meier analysis demonstrated a striking, dose-dependent relationship between the number of elevated metabolites and overall survival (Fig. 2A). Patients with no elevated metabolites had favorable outcomes (median survival not reached at >150 months), whereas those with one, two, three, and four elevated metabolites showed progressively worse survival; patients with all four metabolites elevated had the poorest prognosis (log-rank p<0.0001). The standardized composite score recapitulated this effect: stratification by quartile demonstrated graded survival differences (Fig. 2B; p<0.0001), and a simple median split separated patients into two prognostically distinct groups (Fig. 2C; p<0.0001). These associations remained significant after adjustment for age, sex, and disease site, underscoring the robustness of the composite score as an independent prognostic indicator.

**Fig. 2.**
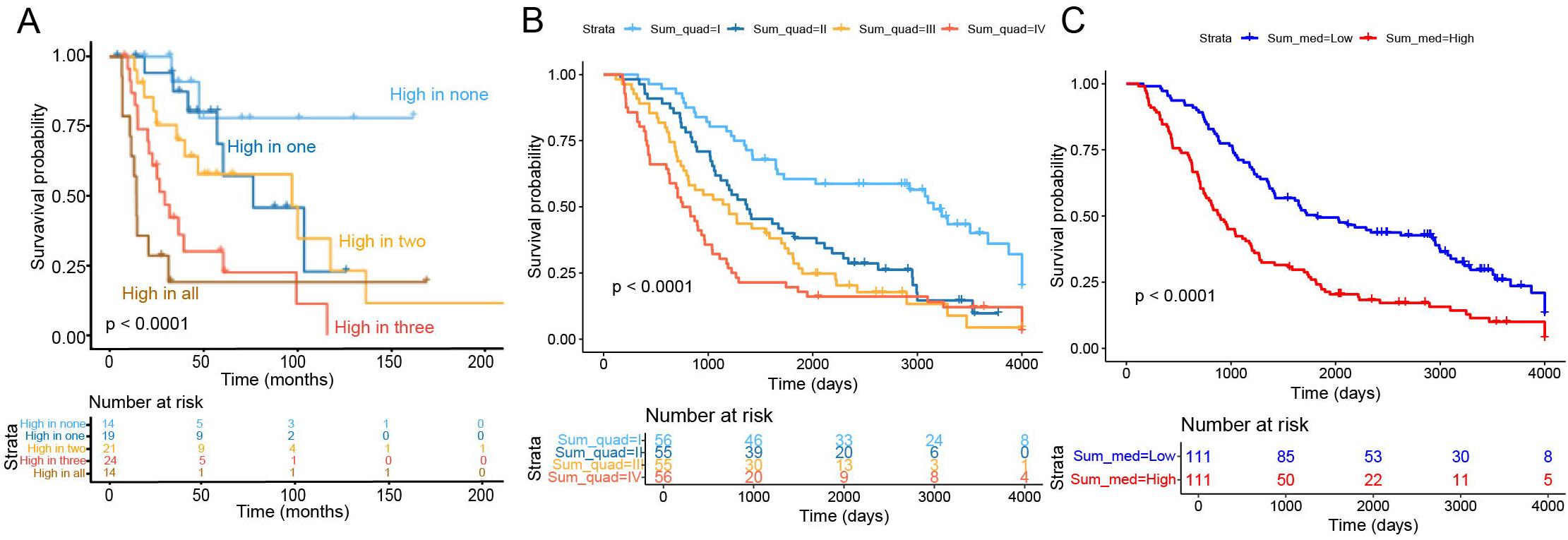
Elevated urinary metabolites are associated with poor overall survival in mesothelioma. (A) Patients were dichotomized at the median for each of the four metabolites and grouped by the number of elevated metabolites (“High in none” through “High in all”); log-rank p<0.0001. (B) Kaplan–Meier survival by quartile of the standardized composite metabolite score (Sum_quad I–IV); log-rank p<0.0001. (C) Kaplan–Meier survival by median split of the composite score (Sum_med Low vs High); log-rank p<0.0001. Risk tables are shown below each panel.

### 3.5 Mechanistic and translational implications

Mechanistically, the four metabolites map onto distinct but cancer-relevant axes. CR has been characterized as a cancer-cell–derived metabolite linked to arginine auxotrophy and altered creatine flux [8]; aberrant sialylation, reflected by elevated NANA, drives immune evasion and metastasis in multiple solid tumors [9]; CS reports on glucocorticoid recycling, a mechanism by which tumors locally generate immunosuppressive steroids to expand regulatory T cells and promote growth [10]; and CP captures dysregulated cholesterol/bile-acid metabolism, increasingly tied to tumor progression. The concordance of the composite score with a 48-gene signature enriched for cell-cycle and DNA-replication programs [7] suggests that these systemic metabolic shifts couple to proliferative tumor states, consistent with the broader concept of metabolic reprogramming as a hallmark of cancer.

Translationally, urine offers clear advantages over blood for serial sampling: it is noninvasive, inexpensive, repeatable, and free from the thrombin-cleavage and clotting variability that complicate fibulin-3 and osteopontin measurements [3,4]. A four-metabolite panel measured by UPLC-MS/MS could complement MESOMARK [2] and existing radiographic assessment, and is well suited to (i) baseline risk stratification, (ii) longitudinal monitoring during therapy, and (iii) enrichment of clinical trials for patients most likely to benefit from intensified or experimental regimens. Integration with transcriptomic or proteomic biomarkers may further refine prognostication.

This study has limitations. It is based on a single-center cohort of moderate size; prospective, multi-institutional validation will be required to establish clinical thresholds and assess performance across histologic and molecular subtypes. Mechanistic studies are needed to determine whether the four metabolites reflect direct tumor secretion, host–tumor metabolic coupling, or systemic adaptation. Standardized, clinically deployable assays—ideally compatible with hospital chemistry workflows—will be necessary for routine translation.

In conclusion, we identify a noninvasive, four-metabolite urinary signature that stratifies survival in malignant mesothelioma and correlates with a validated poor-prognostic molecular phenotype. The dose-dependent relationship between elevated metabolite count and survival, together with the robustness of the composite score, supports its potential clinical utility for risk stratification, monitoring, and trial design in this difficult-to-treat cancer.

## Data Availability

All data produced in the present study are available from the corresponding authors upon reasonable request, in accordance with NIH and Institutional Review Board guidelines.

## CRediT authorship contribution statement

Leila Toulabi: Writing – original draft, Methodology, Investigation, Formal analysis, Data curation, Conceptualization. Joshua K. Stone: Writing – original draft, Visualization, Formal analysis. Mohammed Khan: Visualization, Formal analysis. Daxesh P. Patel: Writing – review & editing, Resources, Methodology, Investigation. Raffit Hassan: Writing – review & editing, Resources, Investigation, Conceptualization. Curtis C. Harris: Writing – review & editing, Supervision, Resources, Project administration, Funding acquisition, Conceptualization.

## Ethics approval and consent to participate

All procedures involving human participants were conducted under an NCI Institutional Review Board–approved protocol (NCT01950572) in accordance with the Declaration of Helsinki and ICH-GCP. Written informed consent was obtained from all participants prior to enrollment.

## Availability of data and material

The dataset analyzed in this study will be made available to qualified researchers upon reasonable request to the corresponding author, subject to institutional data-use agreements and ethical approvals.

## Preprint

An earlier version of this work has been posted on medRxiv: Toulabi L, Stone JK, Khan M, Patel DP, Hassan R, Harris CC. Urinary metabolites and survival in malignant mesothelioma. *medRxiv* 2025.09.23.25336488 (posted 25 September 2025). DOI: https://doi.org/10.1101/2025.09.23.25336488. The preprint is shared in accordance with Elsevier’s article-sharing policy and does not constitute prior publication.

## Acknowledgments

We gratefully acknowledge Betsy Morrow and Jingli Zhang (Thoracic and GI Malignancies Branch, Center for Cancer Research, NCI) and Bhavik Dalal (Laboratory of Human Carcinogenesis, NCI) for expert sample preparation and technical support. We are deeply grateful to the patients and their families who participated in the Mesothelioma Natural History clinical trial (NCT01950572); their contributions made this work possible. The contributions of NIH authors were made as part of their official duties and are Works of the United States Government; the findings and conclusions are those of the authors and do not necessarily reflect the views of the NIH or the U.S. Department of Health and Human Services.

## Supplementary materials

Supplementary material associated with this article (extended Materials and methods; Supplementary Table S1; Supplementary Figs. S1–S3) can be found in the online version.

