## Supplement_Material for "Urinary Metabolite Signatures Stratify Survival in Malignant Mesothelioma: A Noninvasive Prognostic Tool"

**Supplementary Methods**

S1. Patient cohort and study design

S2. Urine collection and storage

S3. Metabolite extraction and UPLC-MS/MS

S4. Gene-expression signature scoring

S5. Composite metabolite score

S6. Statistical analysis

**Supplementary Tables**

Supplementary Table S1. Clinicodemographic characteristics of the mesothelioma cohort (n=95)

**Supplementary Figures**

Supplementary Fig. S1. Urinary metabolite concentrations stratified by age at diagnosis

Supplementary Fig. S2. Urinary metabolite concentrations stratified by disease site

Supplementary Fig. S3. Urinary metabolite concentrations stratified by sex

**Supplementary Notes**

48-gene poor-prognostic signature description

Composite metabolite score and survival-analysis code availability

**References**

References cited in the Supplementary Materials are numbered as in the main text.

**Supplementary methods**

***S1. Patient cohort and study design***

Biospecimens were collected as part of the Tissue Procurement and Natural History Study of Patients with Malignant Mesothelioma (ClinicalTrials.gov: NCT01950572) under an Institutional Review Board–approved protocol of the National Cancer Institute (NCI), and in accordance with the International Conference on Harmonisation–Good Clinical Practice (ICH-GCP) guidelines and the Declaration of Helsinki. All participants provided written informed consent. Between September 2013 and November 2019, 425 patients with histologically confirmed mesothelioma (Board-Certified Anatomic Pathologists, NCI Laboratory of Pathology) were enrolled regardless of asbestos exposure, sex, age at diagnosis, or personal or family history of cancer. RNA-sequencing data were available for 100 patients; the present analysis focused on the 95 patients with matched urine metabolomics and transcriptomic data. Clinicodemographic characteristics of the analyzed cohort are summarized in Supplementary Table S1.

***S2. Urine collection and storage***

Urine samples were collected from study participants, processed locally, and stored at −80 °C until analysis. Samples were thawed on ice immediately prior to extraction. No freeze–thaw cycles beyond one were permitted for analyses reported here.

***S3. Metabolite extraction and UPLC-MS/MS***

Urine extracts were prepared and analyzed by ultraperformance liquid chromatography–tandem mass spectrometry (UPLC-MS/MS) as previously described in Mathé et al. [5] and Parker et al. [8]. Analytical standards were either synthesized in-house or purchased (MilliporeSigma, St. Louis, MO; Cambridge Isotope Laboratories, Tewksbury, MA). Concentrations of creatine riboside (CR), N-acetylneuraminic acid (NANA), cortisol sulfate (CS), and 27-nor-5β-cholestane-3α,7α,12α,24,25-pentol (cholestane pentol, CP) were measured using a Waters XEVO G2 ESI QTOF mass spectrometer. CR (m/z 264.1196, retention time [RT] 0.4 min) and CP (m/z 561.3435, RT 6.3 min) were measured in electrospray ionization (ESI) positive mode; NANA (m/z 308.0982, RT 0.4 min) and CS (m/z 441.162, RT 5.5 min) were measured in ESI negative mode. Concentrations were calculated against calibration curves of analytical standard solutions using MassLynx software (Waters Corporation, Milford, MA) and normalized to urinary creatinine. For dichotomized analyses, patients with metabolite concentrations below the cohort median were classified as “low” and those equal to or greater than the median as “high” (median values: CR 1.987 µM, NANA 9.234 µM, CS 21.011 nM, CP 5.877 nM).

***S4. Gene-expression signature scoring***

A previously validated 48-gene poor-prognostic expression signature for pleural and peritoneal mesothelioma was applied to matched RNA-seq data, as described by Nair et al. [7]. Per-patient signature scores were computed using the published gene list and method, with standardization performed within the cohort to allow comparison across metabolite strata.

***S5. Composite metabolite score***

To integrate information across the four biomarkers, individual metabolite concentrations were z-transformed within the cohort, summed across the four metabolites, and divided by four to yield a per-patient mean z-score (the composite score). Patients were stratified into quartiles (Sum_quad I–IV) and into low/high groups by median split (Sum_med) for survival analyses. The composite score was also evaluated as a continuous variable in correlation analyses against the 48-gene signature.

***S6. Statistical analysis***

All analyses were performed in R 4.1.1. Group comparisons used the Mann–Whitney U test for two-group continuous comparisons; correlations used Spearman’s rank coefficient. Survival was analyzed by the Kaplan–Meier method with log-rank tests using the R packages survminer and survival, and independently verified in Stata. Multivariable Cox proportional-hazards models were used to assess independence of the composite score from age, sex, and disease site. Two-sided p<0.05 was considered statistically significant. No adjustment for multiplicity was applied for the four-metabolite hypothesis-confirming analyses; all reported p-values are nominal.

**Supplementary Table S1**

**Supplementary Table S1. Clinicodemographic characteristics of the mesothelioma cohort (n=95).**

| **Patient characteristics** | **n (%)** |
| --- | --- |
| **Total number of patients** | 95 |
| **Gender** |  |
| Male | 51 (54) |
| Female | 44 (46) |
| **Age at diagnosis (years)** |  |
| <60 | 59 (62) |
| ≥60 | 35 (37) |
| Unknown | 1 (1) |
| **Asbestos exposure*** |  |
| Yes | 52 (55) |
| No | 19 (20) |
| Unknown | 24 (25) |
| **Mesothelioma site** |  |
| Pleura | 47 (49) |
| Peritoneum | 44 (46) |
| Bi-compartmental | 3 (3) |
| Tunica vaginalis | 1 (1) |
| **Smoking status** |  |
| No | 45 (47) |
| Yes | 47 (50) |
| Unknown | 3 (3) |

**Asbestos exposure self-reported.*

**Supplementary Figure legends**

**Supplementary Fig. S1. Urinary metabolite concentrations stratified by age at diagnosis.**

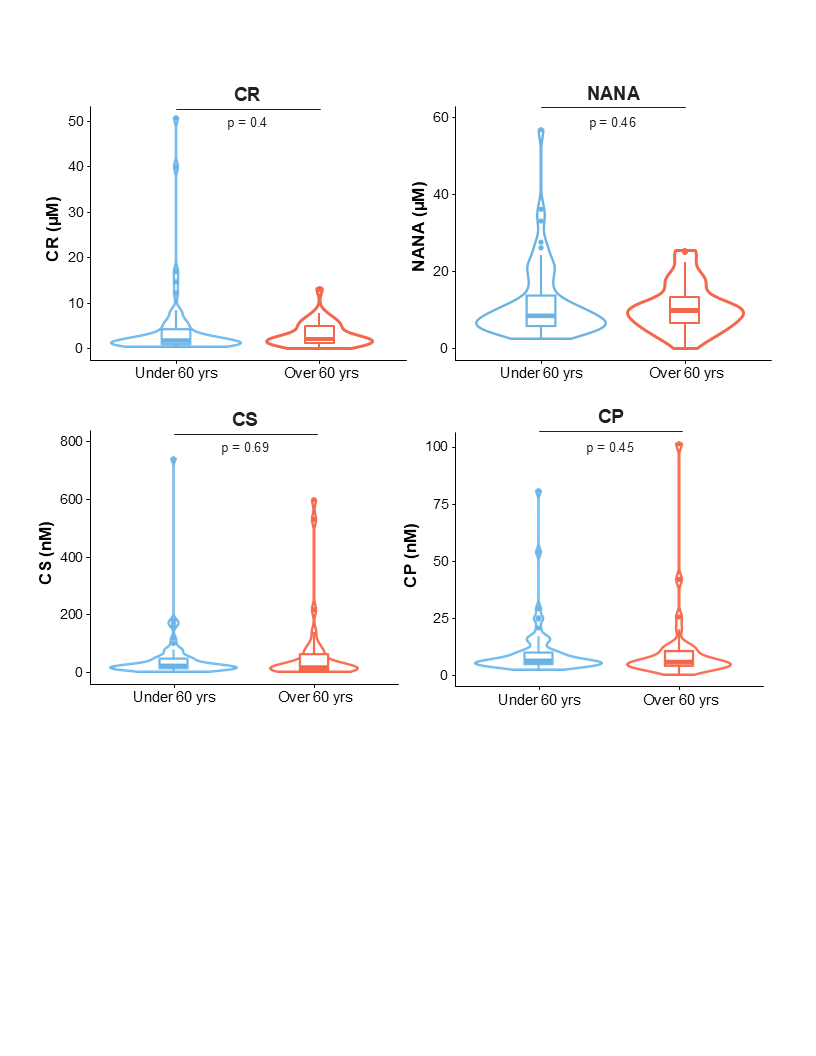

Distribution of urinary CR, NANA, CS, and CP concentrations in patients aged <60 years versus ≥60 years. No significant differences were detected for any metabolite by Mann–Whitney U test. n: CR (<60 n=59, ≥60 n=35); NANA (<60 n=59, ≥60 n=35); CS (<60 n=59, ≥60 n=33); CP (<60 n=59, ≥60 n=34).

**Supplementary Fig. S2. Urinary metabolite concentrations stratified by disease site.**

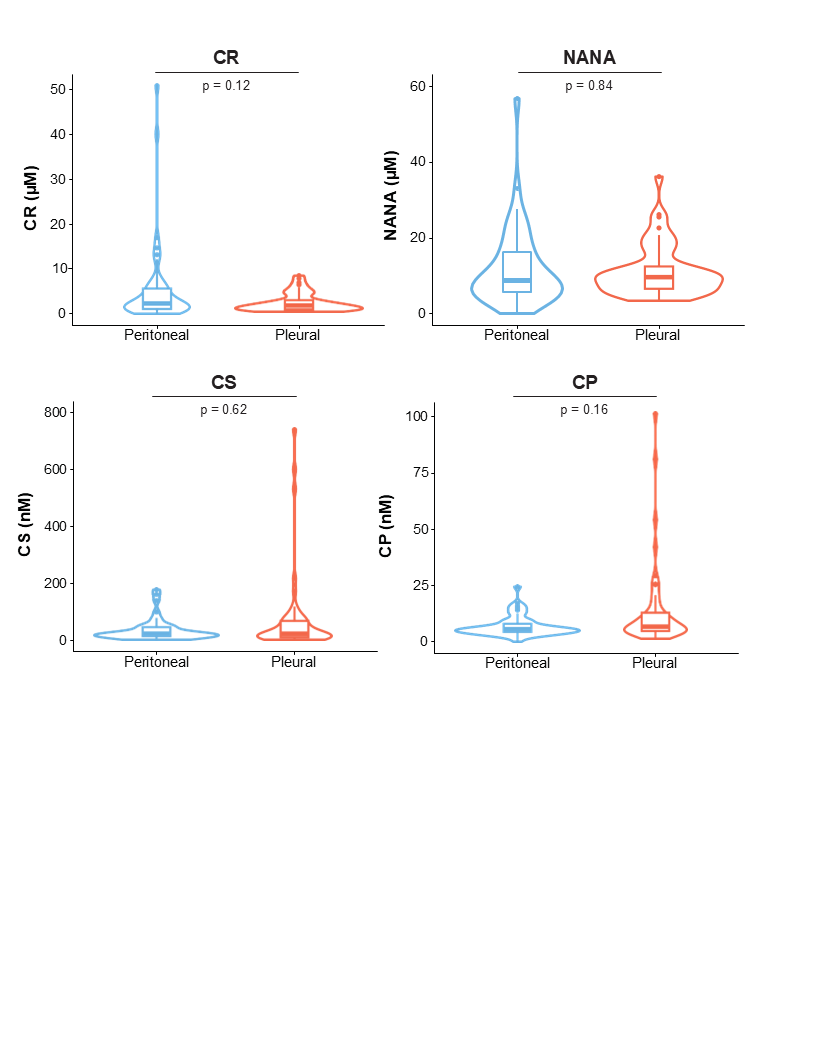
Distribution of urinary CR, NANA, CS, and CP concentrations in patients with peritoneal versus pleural mesothelioma. No significant differences were detected for any metabolite by Mann–Whitney U test. n: CR (peritoneal n=44, pleural n=47); NANA (peritoneal n=44, pleural n=47); CS (peritoneal n=43, pleural n=46); CP (peritoneal n=43, pleural n=47).

**Supplementary Fig. S3. Urinary metabolite concentrations stratified by sex.**

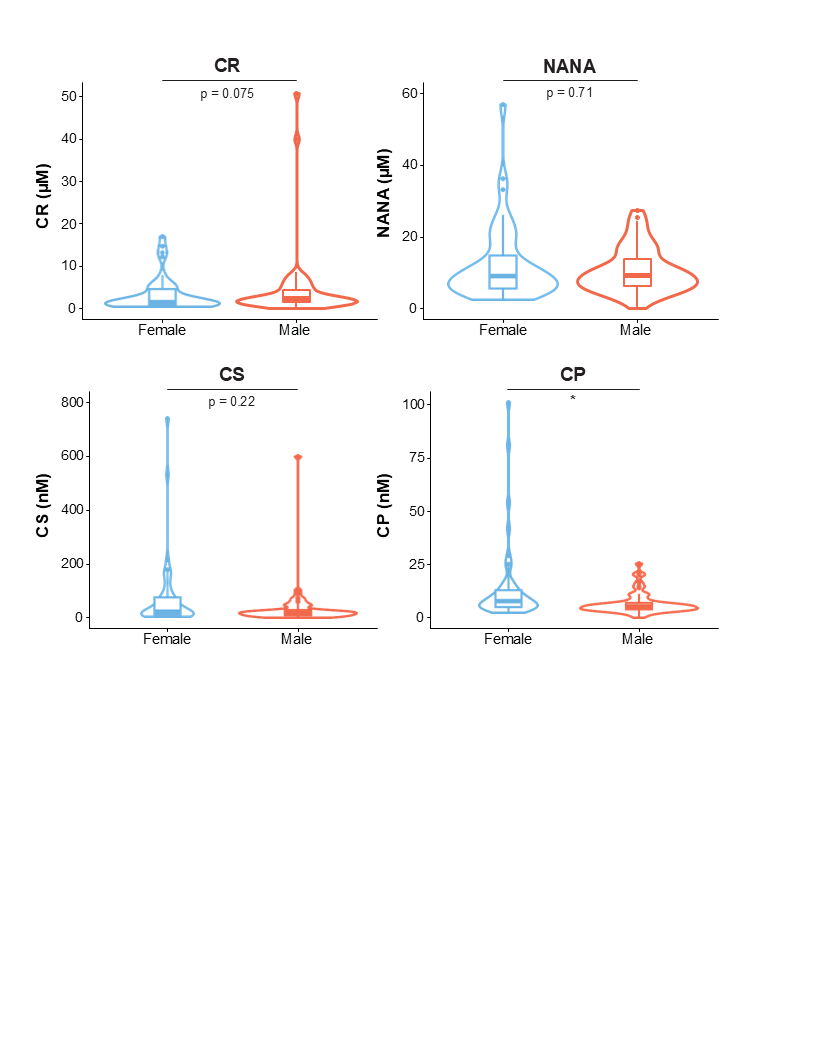

Distribution of urinary CR, NANA, CS, and CP concentrations in female versus male patients. No global sex-based differences were observed for CR, NANA, or CS; CP was modestly higher in female patients. n: CR (female n=44, male n=51); NANA (female n=44, male n=51); CS (female n=44, male n=49); CP (female n=44, male n=50). **p<0.01 by Mann–Whitney U test.

**Supplementary notes**

The 48-gene poor-prognostic signature used in this study was derived and validated in an independent mesothelioma cohort and is enriched for cell-cycle and DNA-replication programs [7]. Per-patient signature scores were computed from RNA-seq normalized counts using the published gene list; higher scores correspond to more aggressive molecular phenotypes. Code used to compute the composite metabolite score and to perform survival analyses is available from the corresponding author upon reasonable request.

**References**

References cited in the Supplementary materials are numbered as in the main text.
